# Implementation and extended evaluation of the Euroimmun Anti-SARS-CoV-2 IgG assay and its contribution to the United Kingdom’s COVID-19 public health response

**DOI:** 10.1101/2021.10.26.21265497

**Authors:** Ashley David Otter, Abbie Bown, Silvia D’Arcangelo, Daniel Bailey, Amanda Semper, Jacqueline Hewson, Matthew Catton, Prem Perumal, Angela Sweed, Jessica Jones, Heli Harvala, Abigail Lamikanra, Maria Zambon, Nick Andrews, Heather Whitaker, Ezra Linley, Alexander J Mentzer, Donal Skelly, Julian C. Knight, Paul Klenerman, PHE Porton Euroimmun testing group, Gayatri Amirthalingam, Stephen Taylor, Cathy Rowe, Richard Vipond, Tim Brooks

**Affiliations:** Diagnostic Support, National Infection Service, Public Health England, Porton, SP4 0JG; SARS-CoV-2 serosurveillance laboratory, National Infection Service, Public Health England, Porton, SP4 0JG; Rare and Imported Pathogens Laboratory, Public Health England, Porton, SP4 0JG; NHS Blood and Transfusion, Microbiology Services, Colindale, UK; Public Health England, Colindale, NW9 5EQ; Seroepidemiology Unit, Public Health England, Manchester, M13 9WZ; Wellcome Centre for Human Genetics, University of Oxford, Oxford, UK; Oxford University Hospitals NHS Foundation Trust, Oxford, UK; Peter Medawar Building for Pathogen Research, Nuffield Department of Medicine, University of Oxford, UK; Nuffield Department of Clinical Neurosciences, University of Oxford, Oxford, UK; National Infection Service, Public Health England, Porton, SP4 0JG; Pathogen Immunology, Public Health England, Porton, SP4 0JG

**Author notes:** Complete list of authors listed in the appendix 1.

## Abstract

In March 2020, the Rare and Imported Pathogens Laboratory at Public Health England, Porton Down, was tasked by the Department of Health and Social Care with setting up a national surveillance laboratory facility to study SARS-CoV-2 antibody responses and population-level sero-surveillance in response to the growing SARS-CoV-2 outbreak. In the following 12 months, the laboratory tested more than 160,000 samples, facilitating a wide range of research and informing PHE, DHSC and UK government policy. Here we describe the implementation and use of the Euroimmun anti-SARS-CoV-2 IgG assay and provide an extended evaluation of its performance. We present a markedly improved sensitivity of 91.39% (≥14 days 92.74%, ≥21 days 93.59%) compared to our small-scale early study, and a specificity of 98.56%. In addition, we detail extended characteristics of the Euroimmun assay: intra- and inter-assay precision, correlation to neutralisation and assay linearity.

## 2. Introduction

Since its emergence in Wuhan, Hubei province, China, in late 2019, the novel severe acute respiratory syndrome coronavirus-2 (SARS-CoV-2) has caused a global pandemic of coronavirus 19 disease (COVID-19). A sobering milestone of three million COVID-19 associated deaths globally was reached on April 16 2021, with a cumulative total of more than 150 million SARS-CoV-2 infections reported worldwide^1^. The true scale of SARS-CoV-2 infections globally is likely to be much higher with a substantial number of cases of mild or asymptomatic disease being undetected.

The UK’s first confirmed case of COVID-19 occurred in a person who had travelled from Hubei province to England on 23 January 2020 and became symptomatic three days later^2^. By the end of February, all four countries of the UK had recorded COVID-19 cases and community transmission was established^3^. Case numbers increased significantly in early March 2020, reaching a total of 6650 laboratory confirmed cases on 23 March^4^ when a national lockdown was implemented. At that time, epidemiological modelling estimated that for every hospitalised COVID-19 case in the UK, there were a further 120-124 infected individuals undetected within the wider community^4,5^.

Individual laboratories began to investigate the use of in-house tests and it became apparent that the UK would benefit from a nationwide service to identify the prevalence of SARS-CoV-2 cases, including asymptomatic ones. Identification of these cases would be essential to inform government policy decisions; to further scientific understanding about immune response evolution, longevity and correlation to protection; and to estimate the true extent of the UK population’s exposure to SARS-CoV-2. Additionally, such a service could address related questions such as the performance of alternative SARS-CoV-2 serology assays. However, this work was complicated by logistical issues including the availability of relevant test kits that were in global demand, and the availability of relevant sample sets to validate new assays.

Described here is the extended evaluation of the SARS-CoV-2 Euroimmun IgG assay, in addition to establishment of a sero-surveillance laboratory using this assay for over 12 months.

## 3. Introduction of an anti-SARS-CoV-2 assay

The Rare and Imported Pathogens Laboratory (RIPL) at PHE Porton Down was tasked by the UK Government’s Department for Health and Social Care (DHSC) with providing a SARS-CoV-2 serology service in support of a national serosurveillance programme. Testing on this scale would require the use of commercial CE-marked serology kits to guarantee the volumes of supply required, the possibility for automation and the necessary quality control of production batches, with our initial focus on an IgG specific assay. An initial market assessment identified only three companies with production kits available. Of these three kits, one targeted antibodies (IgG) against the SARS-CoV-2 spike protein S1 domain, one targeted nucleocapsid, and the third detected antibodies to an unspecified SARS-CoV-2 antigen. Whilst there is merit in having assays that target different antigens for the determination of differential antibody responses to SARS-CoV-2 infection, the use of spike was judged to be critical for virus entry to cells and therefore an assay detecting spike protein antibodies seemed the most attractive for initial use. Thus, the Euroimmun assay (targeting the S1 domain) was selected for the urgent operational requirement and to also provide an indication of the immune profile of subjects.

The laboratory had available existing Stratec Biomedical Gemini platforms (Launch Diagnostics, UK) capable of performing automated ELISAs. After initial ELISA optimisation on the Gemini platform, the first samples were tested to generate initial verification and evaluation data, using samples obtained from patients early in infection and with limited or incomplete clinical data through collaborators/hospitals. On the 30^th^ March 2020, PHE Porton Down went live with a COVID-19 serology service for the UK, with an initial capacity of ∼800 samples a day. This was later expanded upon addition of two further Gemini platforms, doubling capacity to ∼1,600 samples a day. Just over 12 months later, as of 31^st^ of April 2021, over 160,000 samples have been analysed through this assay and workflow, providing answers to key questions about disease spread and the seroprevalence of SARS-CoV-2 infection, as well as informing outbreak management activities. To date, the Euroimmun assay has helped to deliver several significant studies for the UK population, covering population serosurveillance^5^; healthcare worker exposure^6^; selection of donors for convalescent plasma^7–10^; characterisation of reference standards; and research to determine the durability of the immune response^6,11,12^.

## 4. Methods

### Serum samples

Early samples were sourced from collaborating colleagues at PHE Porton Down, PHE Sero-epidemiology Unit Manchester (SEU) and hospitals across England under Material Transfer Agreements (MTAs). These included residual diagnostic sera from SARS-CoV-2 PCR-confirmed samples from hospital in-patients (Guy’s and St Thomas’ Hospital, The Royal Free, Barnet Hospital and North Hampshire County Hospital, Basingstoke and John Radcliffe Hospital, Oxford), with data available on dates of collection versus onset of disease. Samples collected in Oxford were from individuals consented into 2 studies: Gastro-intestinal illness in Oxford: COVID substudy [Sheffield REC, reference: 16/YH/0247] and Sepsis Immunomics project [Oxford REC C, reference:19/SC/0296]) as described^13^. The large majority of samples were sourced from community cases of SARS-CoV-2 described within the FF100^14^ and the PHE serology assay evaluation supporting information^15^. Of the Oxford patient cohort, 116 patients were classified as having mild disease, 15 severe, 13 asymptomatic, 6 critical and 1 death.

To validate the Euroimmun assay (and future serology assays), pre-pandemic negative samples were also sourced to determine specificity. These were obtained from serum banks prior to the COVID-19 outbreak by the Royal College of General Practitioners (RCGP) for routine surveillance schemes and held by PHE SEU.

### Euroimmun anti-SARS-CoV-2 IgG assay

All testing using the Euroimmun anti-SARS-CoV-2 IgG assay (EI 2606-9601 G, Euroimmun, Lübeck) was performed using an automated microplate processor (Stratec Gemini), which automates all aspect of the ELISA from sample dilution, through to incubations, washing, reading and analysis, according to the manufacturer’s instructions for use. Positive and negative kit controls provided by Euroimmun were included with every run, with commercial positive and negative anti-SARS-CoV-2 controls used in parallel (ACCURUN® Anti-SARS-CoV-2 Controls Kit - Series 1000; SeraCare, Gaithersburg) for additional internal quality control. Control values were tracked using a Levy-Jennings plot with Westgard rules applied.

The assay is semi-quantitative, with results expressed as a ratio (index) of the extinction (optical density) of the patient sample (or control) over the extinction of the calibrator. Interpretation of samples were defined according to the manufacturer, with an index value of <0.8 defined as negative, >0.8 to 1.1 defined as indeterminate/borderline and ≥1.1 interpreted as positive.

### Linearity and assay variation

To determine the assay linearity, five high titre serum samples were pooled, with two-fold dilutions generated, and each dilution was run in replicates of five. Similarly, the NIBSC Anti-SARS-CoV-2 Antibody Diagnostic Calibrant (20/162, NIBSC, Potters Bar) was run in triplicate across 1:5, 1:10, 1:20, 1:50 and 1:100 dilutions. Results from both the pool and NIBSC calibrant were used to determine the linearity of the Euroimmun assay.

The pooled serum samples were additionally used to determine the intra- and inter-assay variation, with five replicates measured each day across five days.

### Microneutralisation against SARS-CoV-2

On a panel of 138 convalescent blood donors that were >28 days post-symptom onset, micro-neutralisation titres against SARS-CoV-2 were determined as previously described^7^, with neutralisation titres compared against Euroimmun ratio results.

## 5. Extended verification of the Euroimmun assay

In our original small-scale evaluation of the Euroimmun assay, reported in June 2020^16^ and derived from samples obtained early in the outbreak, 100 PCR-positives samples where the time between sampling and symptom onset was known, were used to determine assay sensitivity, whilst 399 pre-pandemic negatives, 50 serum samples from 2015 seronegative for *Borrelia burgdorferi* and 50 confounder samples (VZV, EBV, CMV and rheumatoid positive) were used to determine specificity (left panel on **Figure 1**). Of the positive panel, 100 samples were initially used but this was later reduced to 93, as seven were found to not be PCR-confirmed. Of the positive samples, 67/93 tested positive (≥1.1 ratio) on the Euroimmun assay, giving an overall sensitivity of 72.0% (95%CI 61.78-80.86) (**Table 1, Figure 3**, PHE Original). When analysing only samples taken ≥14 days post-symptom onset, the sensitivity was 73.42% (95%CI 62.28-82.73) and at ≥21 days, the sensitivity was 74.67% (95%CI 63.30-84.01)^16^.

**Table 1:** Sensitivity of the Euroimmun assay from the original PHE evaluation and the extended evaluation described here. Samples giving an indeterminate value are included with negative results.

|  | All samples |  |  | Samples >14 days |  |  | Samples >21 days |  |  |
| --- | --- | --- | --- | --- | --- | --- | --- | --- | --- |
| Panel | n | sensitivity (%) | 95% CI | n | sensitivity (%) | 95% CI | n | sensitivity (%) | 95% CI |
| Original<sup>16</sup> | 93 | 72.00 | 61.78% to 80.86% | 83 | 73.42 | 62.28% to 82.73% | 77 | 74.67 | 63.30% to 84.01% |
| Extended | 685 | 91.39 | 89.03% to 93.38% | 600 | 92.74 | 90.49% to 94.60% | 579 | 93.59 | 91.41% to 95.36% |

**Figure 1.**
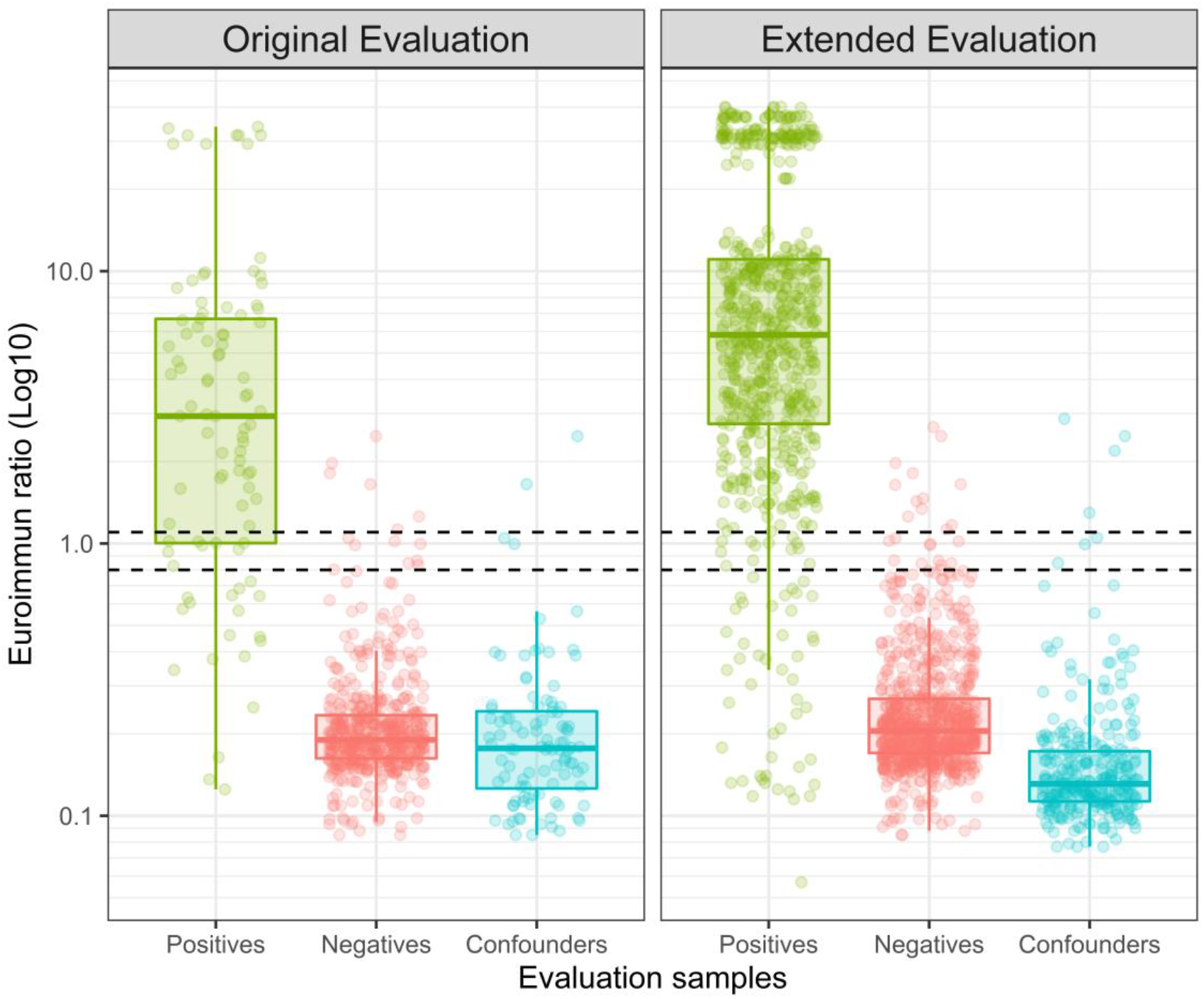
: Distribution of evaluation sample results split according to; **Left**: Original evaluation (n= 692), and **Right**: Extended evaluation (n=1,799). Dashed lines represent the indeterminate range of the Euroimmun assay (ratio or index or s/co ≥0.8 and <1.1).

Subsequent to our initial establishment of the serosurveillance service using the Euroimmun assay, further commercial immunoassay platforms and kits were evaluated at the request of the DHSC to inform decisions on the use of high-throughput assays by NHS laboratories for the detection of anti-SARS-CoV-2 antibodies in patient samples^13,17^. As part of this work, we extended our evaluation of the Euroimmun assay. Due to the limited samples and volumes available for the evaluation of high-throughput serology platforms, the panels differed slightly between our initial Euroimmun evaluation and subsequent other commercial assay evaluations performed by PHE Porton Down^15^. For the extended evaluation, an additional 592 PCR-positive samples that were used for the different verifications of the higher-throughput platforms (including a head-to-head study^13^) were tested using the Euroimmun anti-SARS-CoV-2 IgG assay.

Using this larger panel, the overall sensitivity of the Euroimmun assay was found to be 91.39% (95% CI 89.03% to 93.38%), increasing to 92.74% when using samples with an onset of ≥14 days (95% CI 90.49% to 94.60%) and 93.59% when using samples with an onset of ≥21 days (95% CI 91.41% to 95.36%), markedly improved from the original evaluation (**Figure 1, Table 1, Figure 3**).

Additionally, the specificity panel was expanded from 499 to 1,114 samples: 303 confounders (CMV, VZV, EBV and Rheumatoid positive samples), 711 pre-pandemic negatives and 100 Lyme disease negative samples from 2015. Using this panel, a specificity of 98.56% was calculated (95%CI 97.65-99.17%, n=1,114), similar to that from the original evaluation (specificity of 99.0%; 95%CI, 97.5-99.7%).

Receiver operating characteristic (ROC) analyses showed the highest area under curve (AUC) value was obtained (0.9902) when using samples from the extended evaluation panel with an interval of ≥21 days from onset of symptoms, whilst the original evaluation showed the lowest AUC (0.9584) (**Table 2, Figure 2**).

**Table 2:** ROC analysis values for each of the evaluation panels, with the extended evaluation described here split by total and intervals of ≥14 and ≥21 days.

| Evaluation | Panel | Area | P value | 95% CI | Negatives (incl. confounders) | Positives |
| --- | --- | --- | --- | --- | --- | --- |
| Original<sup>16</sup> | Total | 0.9584 | <0.0001 | 0.9266 to 0.9902 | 499 | 93 |
| Extended | Total | 0.9729 | <0.0001 | 0.9631 to 0.9827 | 1114 | 685 |
| | $\geq 14$ days | 0.9886 | <0.0001 | 0.9826 to 0.9946 | 1114 | 600 |
| | $\geq 21$ days | 0.9902 | <0.0001 | 0.9846 to 0.9958 | 1114 | 579 |

**Figure 2:**
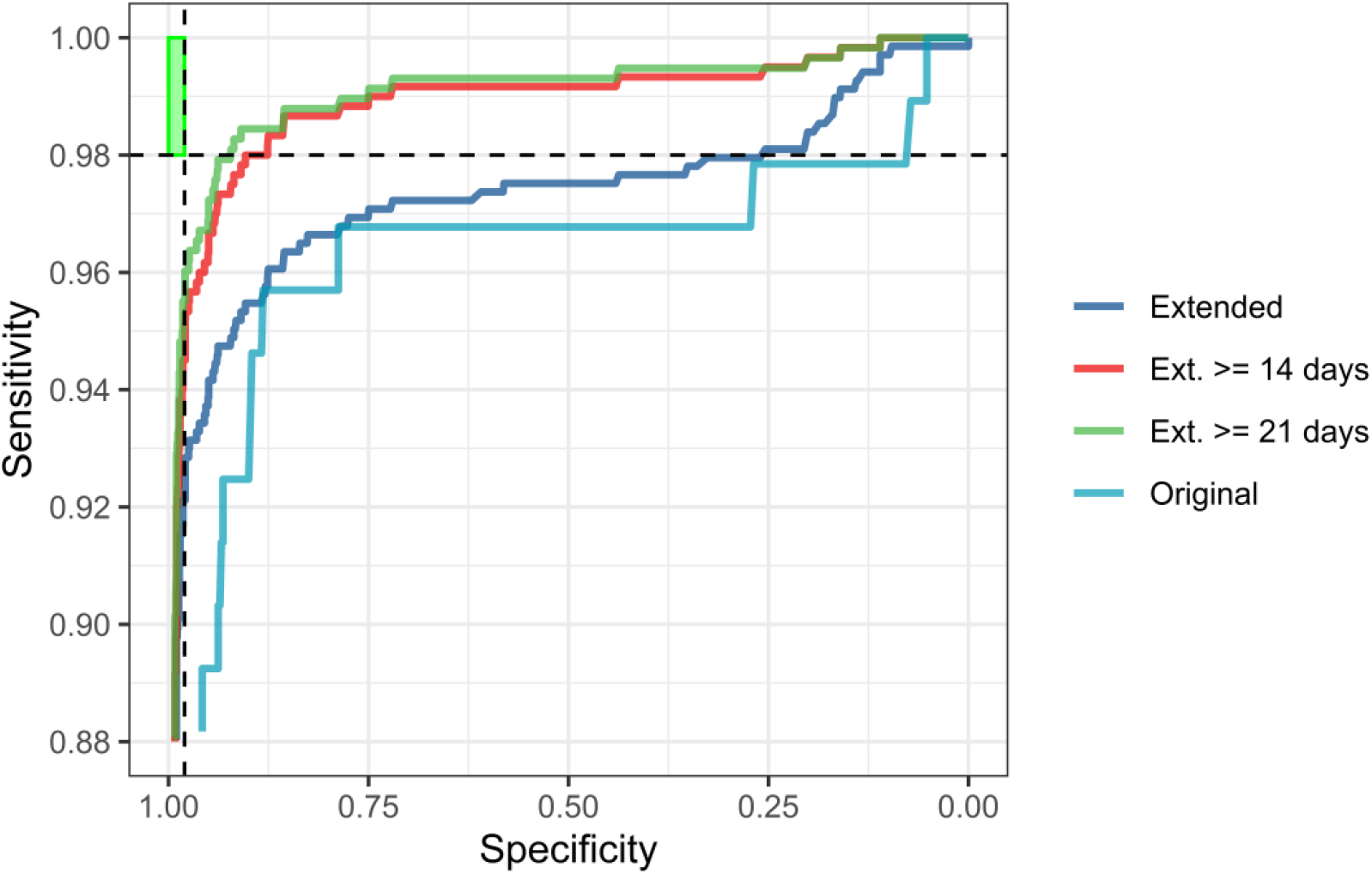
ROC curves for the Euroimmun assay. Curves are shown for the original (n=592) and extended evaluations (n=1,799), with the extended evaluation further split to show samples with intervals of ≥14 (n= 1,714) or ≥21 days (n= 1,693) after symptom onset. The green box indicates the MHRA Target Product Profile of serology assays (>98% sensitivity and specificity) at >20 days.

When examining the temporal distribution of the samples used for each of these evaluations, there was variation in the interval since disease/symptom onset (**Figure 3**). Initial evaluation samples gave the highest frequency of reactive samples at 28 days, whilst the extended evaluation showed the highest interval frequency spanning 36-43 days after symptom onset. The majority of convalescent samples with intervals of <21 days tested positive (25 out of 53 samples).

**Figure 3:**
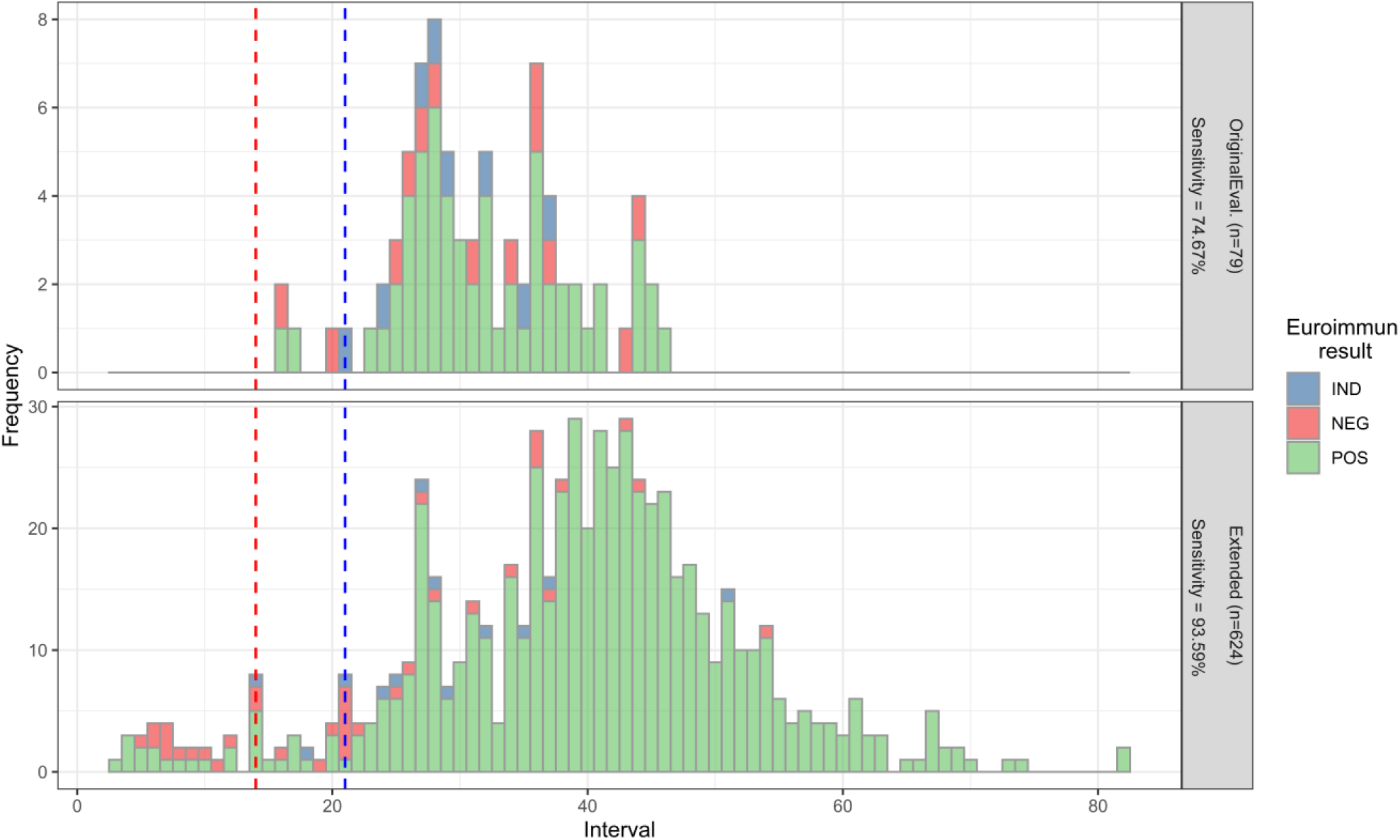
An overview of samples by interval distribution and evaluation panel, coloured by the Euroimmun assay result. Top: Original evaluation (n=79). Bottom: Extended evaluation (n=624). The sensitivity at an interval of ≥21 days is included in each graph, in line with the MHRA TPP. Red dashed line indicates 14-day interval, blue dashed line indicates 21-day interval.

This variation in interval distribution is reflected by the difference in the sensitivities seen between the two evaluations. **Figure 3** clearly shows that many of the initial PHE evaluation samples was limited in interval range, in part by sample availability, with samples typically taken between 14 to 35 days after symptom onset. This may account for differences in sensitivities with some samples potentially taken too soon after onset of symptoms to identify any detectable antibody response. Conversely, many of the extended evaluation samples, by their nature, were taken from convalescent patients who had had sufficient interval post onset to generate a strong antibody response.

However, it should also be noted that no normalisation for sample demographics (for example differences in geographic location, gender, ethnicity or patient age-ranges) had been applied between datasets, so may have occurred and contributed to differences in sensitivity. Analysis of influence of demographic factors between sample sets is beyond the scope of this overview but may be reported in more detail in individual studies. None of our evaluation sets included longitudinal samples from the same patient.

## 6. Further assay characteristics

To determine the precision of the Euroimmun assay, five pooled patient samples were used in the absence of an international standard or calibrant at the time to generate a dilution series for linearity and to determine inter- and intra-assay variation, testing five replicates each over five days. The intra-assay and inter-assay variations were found to be <7% and <5% covariance (CV), respectively, indicating high precision when comparing samples across multiple days and replicate samples (Table S1).

Similarly, the dilution series was used to determine the linear range of the assay, with an R^2^ value of 0.985 (**Figure 4**). As more standards became available, the NIBSC Anti-SARS-CoV-2 Antibody Diagnostic Calibrant was serially diluted to generate additional linearity data, providing a similar R^2^ value of 0.955 (**Figure 4**). From the pooled patient sera and the NIBSC calibrant dilutions, the linear range of the assay was determined to lie between index values of 1 and 9. It is noted that the reader configuration on the Gemini platform resulted in strong positive samples that saturated the OD reader and the software assigns an over-read OD value (e.g. 9.999), which can result in an index value of between 25 – 38 depending on the calibrator OD reading. In routine use, the assay was used in a qualitative mode (e.g. positive/negative interpretation respectively) so saturating values were appropriately assigned as positive.

**Figure 4:**
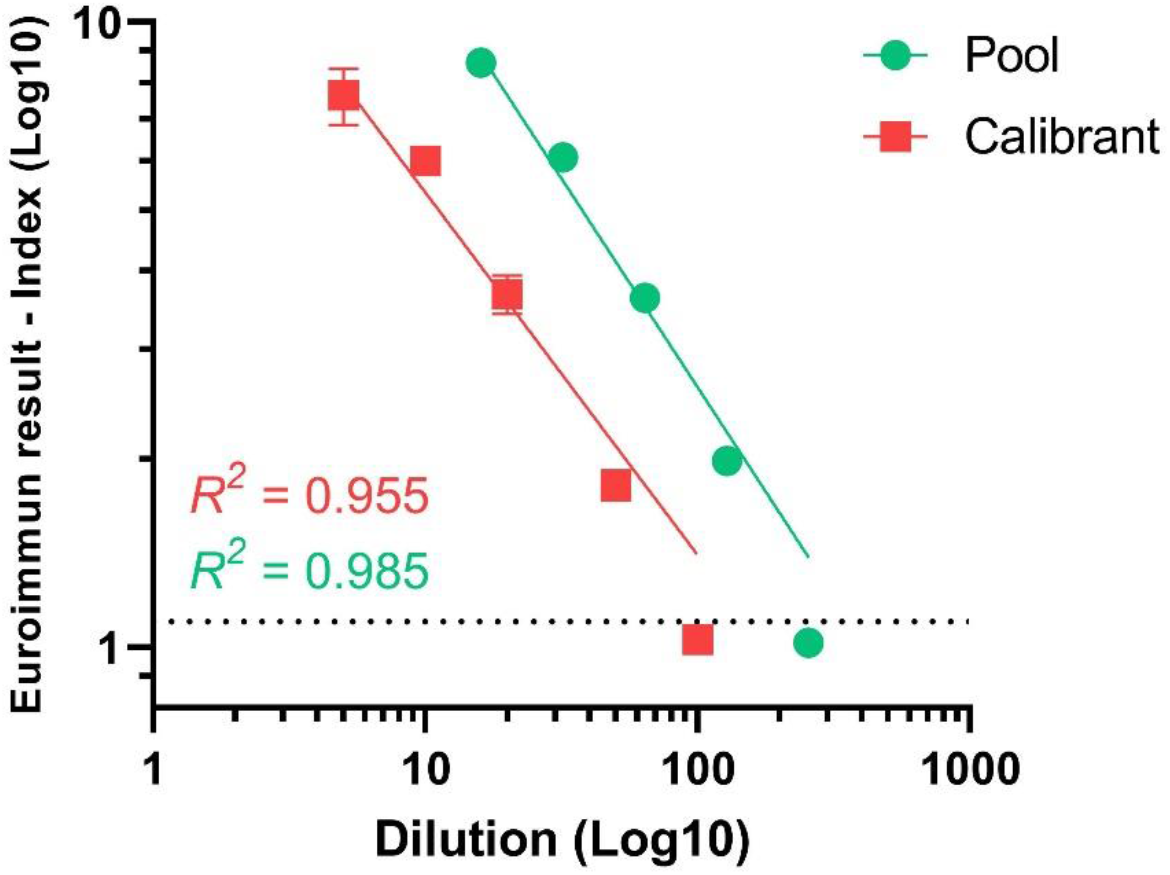
Dilutions and linear range of the Euroimmun assay, using pooled patient samples (pool; green) and the 20/162 NIBSC Anti-SARS-CoV-2 Antibody Diagnostic Calibrant (calibrant; red). Results >9 were removed due to saturated OD readings. Results with an index of ≥1.1 are considered positive whilst those <0.8 are considered negative, with the indeterminate range (0.8 to 1.1) shown with the dashed black line.

Serology assays that have a strong correlation to neutralising antibody titres are useful surrogates for live virus neutralisation assays, which require significant containment level 3 work. Of the 138 samples that were tested by both neutralisation and the Euroimmun assay, 129 were positive on both, whilst nine were only positive by Euroimmun (**Figure 5**), with seven of these samples giving an index result of 1 – 2 (close to the indeterminate range). There was a strong correlation between the Euroimmun results and the neutralisation titre, with an R of 0.83 (R^2^ = 0.69).

**Figure 5:**
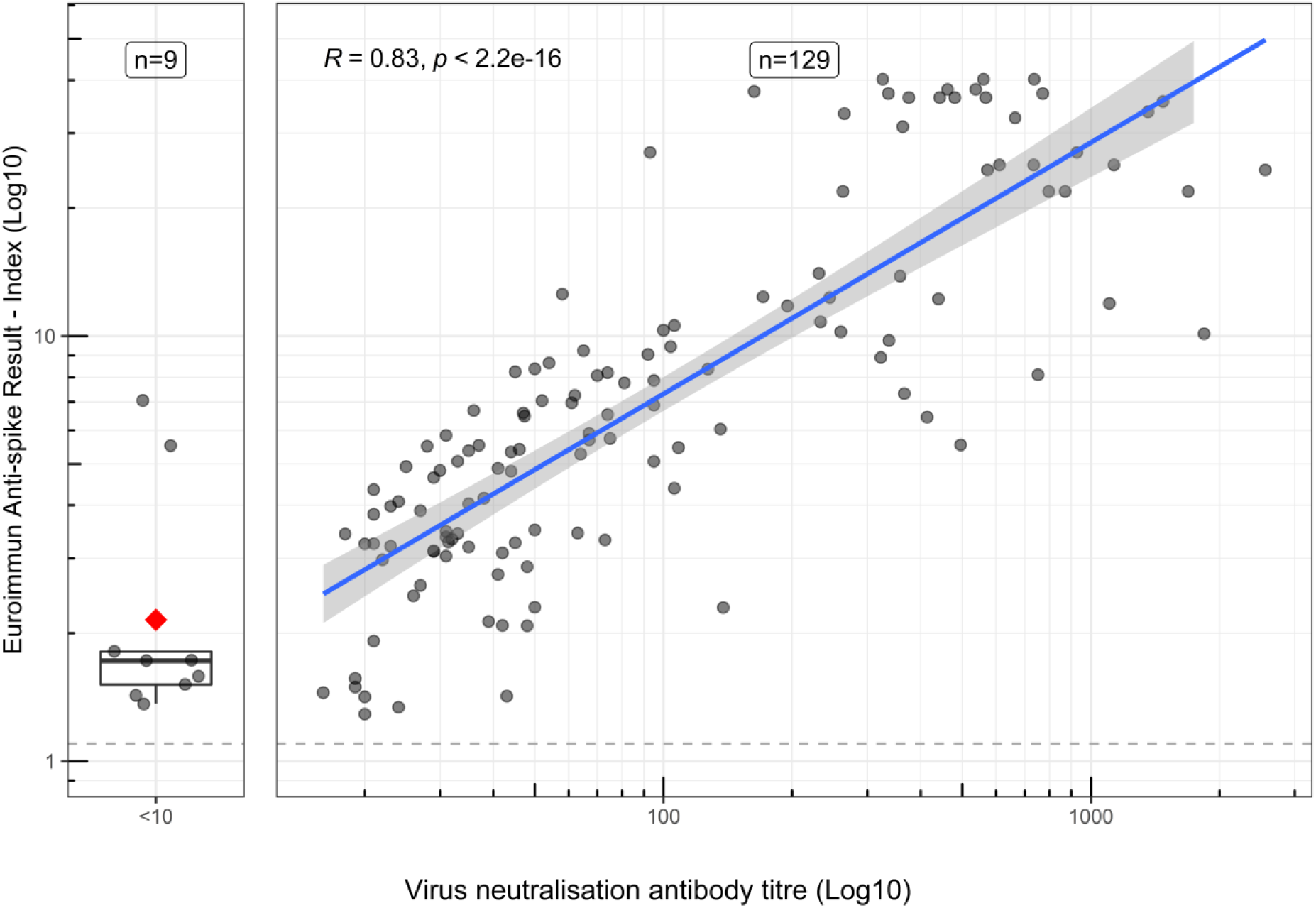
Comparison of convalescent blood donor serum samples tested using the Euroimmun anti-SARS-CoV-2 IgG assay (Log10 ratio value) and virus neutralisation antibody titre (Log10). Good correlation (R=0.83, R^2^=0.69) was observed between neutralisation and the Euroimmun assay. Blue line indicates linear model, with 95% confidence interval (grey range). Dashed line indicates the positive cut-off value (≥1.1 index), whilst the red square shows the mean of the samples negative by neutralisation, but positive by Euroimmun assay.

## 7. Use of the Euroimmun assay in the United Kingdom

A number of national and local projects with appropriate ethical approvals have been facilitated through this work and Figure 6 indicates the scope of Euroimmun testing performed by PHE Porton Down between March 2020 to April 2021. Descriptions of some of these projects are highlighted below although more comprehensive analysis is published elsewhere from the respective project groups.

**Figure 6:**
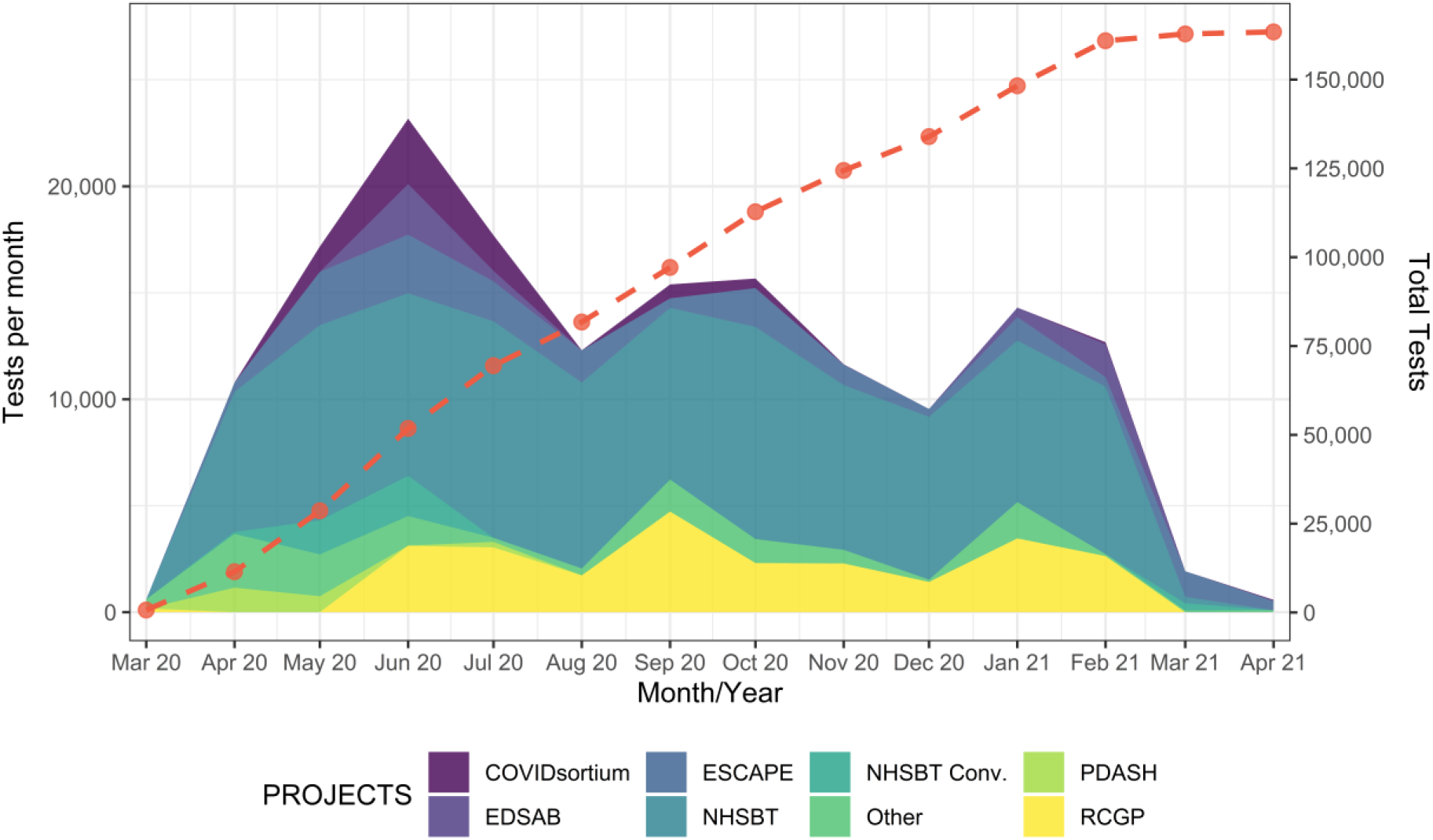
An overview of the number of samples tested using the Euroimmun assay at PHE Porton Down, with total tests (red-dashed line), spanning March 2020 to April 2021. Colours indicate different projects, as described in the text. Projects have since been moved to additional platforms as of March 2021.

As part of a UK serosurveillance scheme, anonymised residual blood donor samples from people aged 17-84 from different regions of England were sourced from the NHS Blood and Transplant (NHSBT) service to provide additional venous blood samples to determine COVID-19 seroprevalence by age and region. This data began prior to other serosurveillance studies (for example, REACT, ONS) so provided PHE and external modelling groups with the only source of seroprevalence data for the UK at the time.

Approximately 10,000 samples were tested monthly and positivity rate on the Euroimmun assay was determined on a weekly basis, to identify the population-level spread of COVID-19 and inform government decision-making. Data was reported to the Public Health England seroepidemiology steering group, scientific pandemic influenza group on modelling (SPI-M) and DHSC, as well as published in the weekly PHE COVID-19 report^5^. This data helped to determine and understand the scale of asymptomatic spread of COVID-19 within the UK population, allowing PHE, DHSC and modellers to assess age groups and areas that are driving transmission.

In addition to the NHSBT serosurveillance, blood samples were obtained from patients visiting primary care settings via the Royal College of General Practitioners (RCGP) Research and Surveillance Centre to determine COVID-19 seroprevalence, with a total of 24,927 samples tested since March 2020.

As part of a collaboration between PHE and NHSBT, convalescent plasma donor samples were tested to determine which donations should be provided for clinical trials assessing the therapeutic effects of convalescent plasma treatment of patients with COVID-19 (NHSBT Conv). The Euroimmun assay was chosen as the results showed a good correlation with neutralisation assays^7,8^; and donations with a Euroimmun serology index result of >9.1 (later revised to 6.0) were shown to be associated with the presence of high neutralising antibody titres of 1:100 or higher^7,8^ and were hence selected for trial use. Serological testing in support of this project began in April 2020, with the first patients receiving treatment in the beginning of May as part of the REMAP-CAP trial^10^ and subsequently via the RECOVERY trial^9^.

Whilst laboratory antibody testing using ELISAs is considered the gold-standard for determining population-level exposure to SARS-CoV-2, additional studies were conducted to determine the feasibility of using lateral flow device antibody tests. On request from DHSC, a study was performed on emergency service workers to evaluate the utility of home antibody testing kits; EDSAB-HOME (Evaluating Detection of SARS-CoV-2 AntiBodies at HOME). Capillary blood samples from >2,500 volunteers were tested on different lateral flow devices and a contemporaneous venous sample was tested on the Euroimmun assay as the reference test^18^.

From the start of the first COVID-19 lockdown in the UK (23 March 2020), the COVIDsortium study (NCT04318314) collected serial samples from a cohort of 731 healthcare workers (HCW) with the aim of identifying host and pathogen correlates of protection and pathogenesis in mild/asymptomatic SARS-CoV-2 infection^19,20^. The Euroimmun assay was used throughout to characterise the serostatus of all study samples which by completion of the final follow-up samples at 1 year, will number more than 13,000 samples. This data helped to define the proportion of this HCW cohort who seroconverted, and was used to monitor the longitudinal antibody response and rate of decline of anti-S1 IgG antibodies^6^ and to investigate the relationship between peak anti-S1 IgG titre and neutralising antibody titre^21^.

In addition, a small project (ESCAPE) began in March 2020 as a mechanism to study antibody responses to infection. A cohort of Public Health England staff across 4 sites were recruited, with monthly blood donations taken as a way to understand the kinetics of antibodies post-infection^11^.

A collaboration between PHE and Hampshire Hospitals NHS Foundation Trust (HHFT), provided 1885 serial samples from 282 hospital in-patients with acute COVID-19 and 189 serial samples from 50 convalescent HCWs (the PDASH study). All participants had PCR confirmed SARS-CoV-2 infections that ranged from asymptomatic to severe disease. The study used the Euroimmun assay to determine the time to seroconversion and to monitor the longitudinal antibody response in study participants.

Other projects included those used specifically for serology assay evaluations conducted by PHE Porton and PHE SEU, smaller research studies, and those conducted by hospitals and colleagues early in the pandemic.

## 8. Discussion

The Euroimmun assay has been fundamental to the UK’s public health response to COVID-19, with >160,000 samples tested March 2020 to March 2021 (**Figure 6**). This early availability of the assay was key to performing seroprevalence testing in the early stages of the pandemic, informing the policy and legislative responses of PHE, DHSC and the UK Government. It also contributed to our understanding of immune responses during COVID-19 infection and recovery as well as provided real-time (∼24 hours) testing of convalescent plasma before being provided for use in clinical trials.

Whilst many platforms and assays now exist for the detection of anti-SARS-CoV-2 antibodies (including those evaluated by ourselves^22^), there were limited choices available at the onset of the pandemic in the UK in March 2020. This highlights the need and basis for pandemic preparedness from an immunology and serology perspective, with the requirement for labs to be able to run high-throughput antibody tests on specimens from patients infected with newly emerging pathogens. Rapid immunology research on a new pathogen enables early understanding of potential immunity and inform early vaccine design. Whilst in-house/research-based assays were available since January 2020, these require significant quality control to ensure consistent diagnosis across batches of samples to ensure consistent sensitivity. Similarly, in-house assays can also suffer from logistical and supply issues for specially manufactured components (recombinant proteins etc). Commercial assays became rapidly available with the spread of SARS-CoV-2 in early 2020, however supply and logistical issues were also evident with the global spread of SARS-CoV-2, so this should be a factor in deciding on assays for serosurveillance.

In our original evaluation, we found an overall sensitivity of 72.0% (interval ≥14 days: 73.42%, interval ≥21 days: 74.67%) and a specificity of 99.0% for the Euroimmun assay, whilst the extended evaluation detailed here found an overall sensitivity of 91.39% (interval ≥14 days: 92.74%, interval ≥21 days: 93.59%) and a specificity of 98.56% (**Table 1, Figure 2**). This data is supported by other studies, which have shown similar high sensitivities of 90%^23^, 96%^24^ and 100%^25^. The initial evaluation suggested a moderate sensitivity for the Euroimmun assay, but this is likely a reflection of the early stages of the pandemic, when serum from patients with confirmed COVID-19 were extremely difficult to obtain, disease onset was poorly characterised and little convalescent material was available. This limited the number of samples, as many of the original evaluation samples were obtained from patients with mild disease within the community, so in using these particular samples, this could have resulted in lower sensitivity than anticipated, due to mild infections and resulting in lower antibody responses than those with severe infections or those requiring hospitalisation^26^. In addition, timing between disease onset and sample date was not fully established for all patient samples, with many taken from patients too early since disease onset, whereby no robust IgG/immunological response is mounted.

Use of an extended sample panel has enabled us to better evaluate the Euroimmun assay. Although the Euroimmun assay showed improved sensitivity in our extended evaluation, in our analysis, it still does not meet the MHRA’s Target Product Profile (TPP) of >98% sensitivity recommendation for SARS-CoV-2 serology assays to determine if people have recent infection of SARS-CoV-2^27^; it does however meet the TPP of >98% specificity.

In addition to improved sensitivity using this extended panel, intra- and inter-assay variation was found to be low, with covariance of <7% and <5%, respectively. Using pooled patient samples and the NIBSC diagnostic calibrant, the linear range of the assay was also determined, enabling semi-quantitative comparisons (**Figure 4**). Our results are consistent with other studies that show that only a defined range of the Euroimmun assay can be used to reliably infer a linear range^6^.

As an anti-spike (S1 domain) antibody ELISA, the Euroimmun assay results also correlate with neutralising antibody titres (R=0.83, R^2^=0.69) as shown elsewhere^7,8,28^, suggesting that this assay provides a useful surrogate for virus neutralisation and possible correlation to immune protection (**Figure 5**), without the requirement for containment level 3 facilities and live-virus culture. However, other pseudo-type assays have shown higher correlation^28^, likely due to the different dynamics between antibody-binding assays and neutralisation-based assays.

In addition, as an indirect sandwich ELISA, results are only semi-quantitative and display a maximum reading for optical densities due to reader saturation. In our equipment implementation depending on the calibrator value, max OD values can then result in an index value of between approximately 26 and 38, preventing direct comparisons between samples unless the calibrator values are similar or are within the linear range of the assay, determined here to be between OD values of 1 and 9, (**Figure 4**). If quantitative results are desired, our recommendation is to dilute samples that are strongly positive to within this range.

Notably, of the serology assays evaluated by PHE Porton Down to date^22^, the Euroimmun assay format does not require the installation of dedicated machinery/specialist equipment, similar to other plate-based ELISA assays. This enables wider use of the assay: it can be utilised in most laboratory settings using automated pipetting platforms (such as the Stratec Gemini), or on the bench with manual pipetting, or within the field since only limited additional laboratory equipment such as a plate reader is required.

Serology assay selection for different requirements is complex^29^. This initial programme was principally focused on serosurveillance, further work is now underway at PHE Porton to determine antibody responses induced by vaccination^30^, vaccine effectiveness^31^, vaccine coverage, variant infection and population spread^32^, using a combination of serology assays targeting different antigens. With additional assays now available, we have expanded our assays to encompass those with improved sensitivity and specificity, with the capability to change rapidly, enabling better and more accurate analysis of the differing and refined responses to the SARS-CoV-2. Newer assays have enabled us to determine specific protein sub-domain responses (e.g. anti-RBD), as well as using a combination of assays as a mechanism to differentiate between those with previous infection (presence of both anti-spike and anti-nucleocapsid antibodies), those with vaccination and no previous infection (presence of only anti-spike antibodies) and those with previous infection and vaccination (presence of both anti-spike and anti-nucleocapsid antibodies, with significantly higher anti-spike responses as reported elsewhere^30^).

## Data Availability

All data produced in the present work are available upon reasonable request to the authors.

## 9. Acknowledgements and funding

We thank the many scientists and clinicians that provided PHE Porton with early samples from those with PCR-confirmed SARS-CoV-2 infection in March 2020, including those at Guy’s and St Thomas’ Hospital, Westminster, Royal Free Hospital, Barnet and North Hampshire County Hospital, Basingstoke. In addition, we thank collaborators from the head-to-head study at Oxford University (Gillian Rodger, Hayleah Pickford, Teresa Street, Kate Dingle, Justine Rudkin, Derrick Crook, David Eyre, Philippa Matthews and Nicole Stoesser).

This project was funded as part of the DHSC serological assay evaluation procedure. AJM and DTS are funded by ISARIC-4C “Medical Research Council (grant MC_PC_19059) as part of the ISARIC Coronavirus Clinical Characterisation Consortium (ISARIC-4C)”, COMBAT Consortium, OUH clinical and research staff and patients

## 10. Author contributions

ADO and AB wrote the manuscript. AB, DB, ADO, SD’A, TB, AS, MC, PP, AS and RV performed and oversaw the initial evaluation and assay implementation. ADO, AB and SD’A performed the extended assay characterisation. AB, ST, JH, CR and SD’A were responsible for routine testing. EL provided initial evaluation samples and EL & AS facilitated with sourcing samples. AJM and DTS provided additional evaluation samples. AL, HH and MZ were responsible for the microneutralisation work. All authors read and commented on the final manuscript.

---

**Table S1:** Precision results for the Euroimmun assay using five pooled patient sera. Samples were tested in replicates of five over five days. Intra-assay variation was <7%, whilst inter-assay variation was <5%

| Sample |  | Day 1 | Day 2 | Day 3 | Day 4 | Day 5 | Inter-assay Mean | Inter-assay SD | Inter-assay CV |
| --- | --- | --- | --- | --- | --- | --- | --- | --- | --- |
| 1 | Mean | 9.00 | 8.43 | 8.59 | 8.43 | 8.64 | 8.62 | 0.235 | 2.724 |
|  | SD | 0.26 | 0.13 | 0.14 | 0.08 | 0.13 |  |  |  |
|  | CV | 2.93 | 1.59 | 1.60 | 0.99 | 1.50 |  |  |  |
| 2 | Mean | 6.28 | 5.95 | 5.99 | 6.04 | 6.12 | 6.08 | 0.132 | 2.180 |
|  | SD | 0.11 | 0.19 | 0.13 | 0.16 | 0.28 |  |  |  |
|  | CV | 1.69 | 3.26 | 2.18 | 2.64 | 4.50 |  |  |  |
| 3 | Mean | 3.72 | 3.52 | 3.55 | 3.67 | 3.67 | 3.62 | 0.087 | 2.409 |
|  | SD | 0.11 | 0.12 | 0.04 | 0.19 | 0.11 |  |  |  |
|  | CV | 2.84 | 3.52 | 1.13 | 5.17 | 2.93 |  |  |  |
| 4 | Mean | 2.07 | 1.90 | 1.99 | 1.99 | 1.99 | 1.99 | 0.062 | 3.102 |
|  | SD | 0.04 | 0.03 | 0.06 | 0.09 | 0.08 |  |  |  |
|  | CV | 2.16 | 1.72 | 3.12 | 4.42 | 4.04 |  |  |  |
| 5 | Mean | 1.07 | 0.98 | 1.02 | 0.99 | 1.03 | 1.02 | 0.037 | 3.620 |
|  | SD | 0.07 | 0.01 | 0.04 | 0.02 | 0.07 |  |  |  |
|  | CV | 6.28 | 0.55 | 3.76 | 2.46 | 6.60 |  |  |  |

**Table S2:**
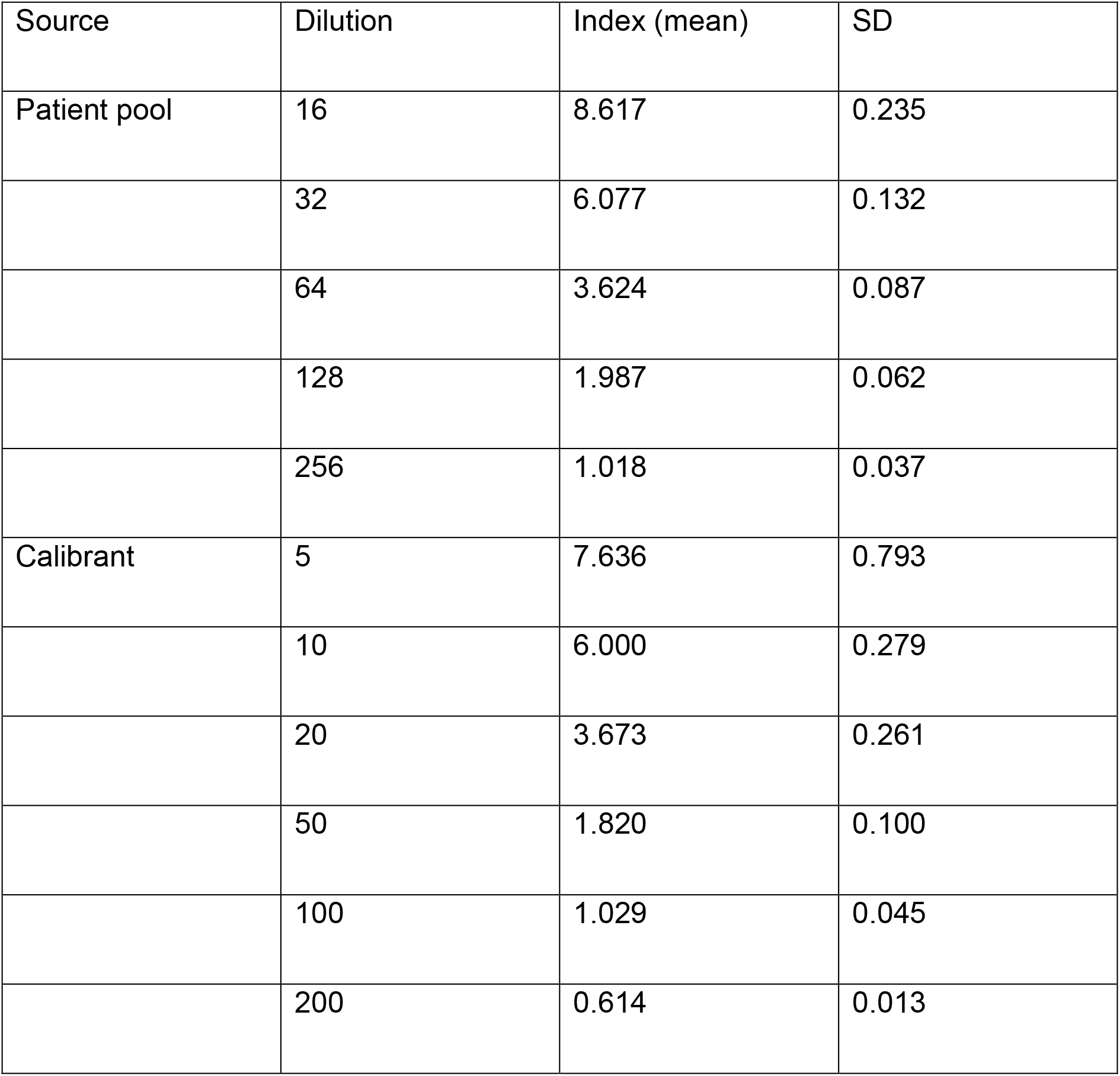
Determination of linear range of EuroImmun assay using serially diluted pooled patient sera. Pooled serum samples at dilutions of 2, 4 and 8 were removed due to OD saturation.

## 12. Appendix 1

### PHE Porton Euroimmun testing group list of authors

Joanna Bacon, Daniel Bailey, Natalie Baker, Jack Bleach, Debbie Blick, Paul Bolton, Abbie Bown, Olivia Carr, Matthew Catton, Melanie Clifford, Tom Coleman, Charlie Cooper, Charlotte Cornbill, Ellie Cross, Mollie Curran-French, Silvia D’Arcangelo, Owen Daykin-Pont, Charlotte Dixon, Phoebe Do Carmo Silva, Ellie Drinkwater, Jackie Duggan, Xena Dyball, Jack Easton, Ruth Elderfield, Anaya Ellis, Jenna Furneaux, Harriet Garlant, Barry Gibney, Supriya Gurung, Rachel Halkerston, Robin Hanson, James Hardy, Alexander Hargreaves, Gabrielle Harker, Becky Harrison, Bethany Hicks, Charlotte Hind, Emma Hobbs, James Hogg, Margarita Jablonska, Leah Johal, Caoimhe Kelly, Carensa Knights, Emily Laing, Deborah Lister, Rosa Loffredo, Adam Mabbutt, Bethany Martin, Felicity Marx, Lara Mason, Joanna McGlashan, Debbie McKee, Isobel Miles, Sebastian Millward, Gloria Mongelli, Christopher Moon, Taalia Morgan, Alexandra Morrison, Alexandra Morrison, Anna Moy, Josh Nelthorpe-Cowne, Jack Nevols, Jane Osbourne, Jodie Owen, Ros Packer, Jordan Pascoe, Prem Perumal, Esther Picton-Barlow, Eleanor Porges, Saskia Proud, Deen Qureshi, Anastasia Richardson, April Roberts, Adam Roberts, Amanda Semper, Lauren Setterfield, Jennifer Small, Kirstie Speer, Sara Speight, Angela Sweed, Stephen Taylor, Steve Thomas, Sian Tiley, Elizabeth Truelove, Ann Varghese, Richard Vipond, Rosie Watts, Katie West, Clare Wilson, Kate Withycombe, Charlotte Woolley.

